# Lipid-based nutrient supplements for prevention of child undernutrition: when less may be more

**DOI:** 10.1101/2023.06.28.23292006

**Authors:** Kathryn G. Dewey, Charles D. Arnold, K. Ryan Wessells, Christine P. Stewart

**Affiliations:** Institute for Global Nutrition and Department of Nutrition, University of California, Davis, Davis, CA, USA (KGD, CDA, KRW, CPS)

**Keywords:** malnutrition, infants, home fortification, food supplements, linear growth

## Abstract

**Background:** Both small-quantity (SQ) and medium-quantity (MQ) lipid-based nutrient supplements (LNS) have been used for prevention of child undernutrition. A meta-analysis of 14 trials of SQ-LNS vs no LNS showed effects on length-for-age (LAZ, +0.14 (95% confidence interval 0.11, 0.16)) and weight-for-length (WLZ, +0.08 (0.06, 0.10)) z-scores, as well as prevalence ratios (95% CI) for stunting (LAZ<-2, 0.88 (0.85, 0.91)) and wasting (WLZ < −2, (0.80, 0.93)). However, little is known about the effects of MQ-LNS on growth.

**Objective:** We aimed to examine the effects of preventive MQ-LNS (∼250-499 kcal/d) provided at ∼6-23 mo of age on growth outcomes compared to no LNS or provision of SQ-LNS.

**Methods:** We conducted a systematic review of studies of MQ-LNS for prevention, and categorized them as providing < 6 mo vs. ≥ 6 mo of supplementation; for the latter category we conducted a meta-analysis, with main outcomes being change in WLZ and LAZ, and prevalence of wasting and stunting.

**Results:** Three studies provided MQ-LNS for 3-5 mo (seasonal) for children 6-36 mo of age, and did not show consistent effects on growth outcomes. Eight studies provided MQ-LNS for 6-18 mo, generally starting at 6 mo of age; in the meta-analysis (max total n=13,673), MQ-LNS increased WLZ (+0.09 (0.05, 0.13)) and reduced wasting (0.89 (0.81, 0.97)), but had no effect on LAZ (+0.04 (−0.02, 0.11)) or stunting (0.97 (0.92, 1.02)) compared to no LNS. Two studies directly compared SQ-LNS and MQ-LNS and showed no significant differences in growth outcomes.

**Conclusions:** The current evidence suggests that MQ-LNS offers no added benefits over SQ-LNS, although further studies directly comparing MQ-LNS vs. SQ-LNS would be useful. One possible explanation is incomplete consumption of the MQ-LNS ration and thus lower than desirable intake of certain nutrients.

**Registry:** PROSPERO CRD42022382448: https://www.crd.york.ac.uk/prospero/display_record.php?ID=CRD42022382448

## Introduction

Lipid-based nutrient supplements (LNS) provide multiple micronutrients embedded in a food base that also provides energy, protein and essential fatty acids (1). There is a wide range in the size of the daily ration of LNS provided to young children, depending on whether they are aimed at prevention or treatment of undernutrition (2). The full suite of LNS products includes small-quantity (SQ-LNS, typically 100-120 kcal/d), medium-quantity (MQ-LNS, typically 250-499 kcal/d) and large-quantity (LQ-LNS, typically > 500 kcal/d) LNS. The most widely known product is Ready-to-Use Therapeutic Food (RUTF), which is a type of LQ-LNS used for treatment of severe wasting, with the dosage usually based on body weight and treatment goal. On the other end of the spectrum, SQ-LNS are designed for prevention, not treatment, of undernutrition among children 6-23 mo of age. MQ-LNS have been used for both prevention and treatment, and the quantities provided have varied considerably. The ration size of MQ-LNS currently used by the World Food Programme (WFP) and several other agencies is 50 g/d (255 kcal/d), generally aimed at prevention of seasonal wasting and/or child undernutrition in food-insecure populations (3).

The evidence for preventive effects of SQ-LNS on stunting, wasting, iron-deficiency anemia and mortality is based on meta-analyses of 14-18 randomized controlled trials that included >37,000 children 6-23 mo of age in low- and middle-income countries (4–6). This evidence base was rated as “strong” in the 2021 Lancet Series on Maternal and Child Undernutrition (7). By contrast, less is known about the impact of MQ-LNS used for prevention. In a Cochrane review in 2019, Das et al. (8) found no significant differences in growth outcomes between SQ-LNS and MQ-LNS for children 6-23 mo of age when results were stratified by LNS quantity, but data were available for only 4 MQ-LNS trials.

This previous evidence thus raised the question of whether there is any advantage of MQ-LNS over SQ-LNS for prevention of undernutrition among children 6-23 mo of age. Given that MQ-LNS is more expensive per daily ration than SQ-LNS (9, 10), answering this question has important programmatic implications. Some studies have suggested that children of this age, especially those < 12 mo of age, are unlikely to consume the full daily ration of MQ-LNS in addition to breast milk and other complementary foods provided by caregivers (11–13). The energy needed from complementary foods, assuming average breast-milk intakes, is only ∼200 kcal at 6–8 mo, ∼300 kcal at 9–11 mo, and ∼550 kcal at 12–23 mo of age (14). A daily ration of 255 kcal/d from MQ-LNS exceeds or comprises a large percentage of those targets. Thus, when given MQ-LNS, the child may leave a substantial amount of the supplement unconsumed. In Malawi, for example, when MQ-LNS providing 241 kcal/d was provided and the caregiver reported LNS consumption on the day of dietary assessment, average intake of LNS was only ∼100 kcal/d at ∼9 mo of age (13), meaning that those infants received less than half of the intended amounts of the vitamins and minerals. In Pakistan, only 20.7% of children consumed the full dose of MQ-LNS when offered, even though 81% of caregivers were aware of the recommended dose (12).

To provide updated evidence relevant to this question, we aimed to examine the effects of preventive MQ-LNS provided at ∼6-23 mo of age on child growth outcomes compared to (1) no provision of LNS and (2) provision of SQ-LNS. To understand the implications of incomplete consumption of the daily ration of MQ-LNS, we also estimated nutrient intakes from MQ-LNS and SQ-LNS under different scenarios.

## Methods

### Systematic review and meta-analysis

The protocol for this review and meta-analysis was registered as PROSPERO 2022 CRD42022382448, https://www.crd.york.ac.uk/prospero/display_record.php?ID=CRD42022382448. The protocol was originally registered on December 18, 2022 and modified on May 19, 2023 to clarify the definition of the control group. We have reported results according to the Preferred Reporting Items for Systematic Reviews and Meta-Analyses (PRISMA) guidelines (15). A detailed protocol is available on Open Science Framework (16).

### Literature search and criteria for inclusion in meta-analysis

We began the search by considering the studies identified by and included in a 2019 Cochrane systematic review and meta-analysis of the provision of preventive LNS for children 6-23 mo of age (8), as well as a series of individual participant data (IPD) analyses that examined the effects of SQ-LNS on the prevention of child undernutrition and promotion of healthy development (5). Then we repeated the database search methods employed in the 2019 Cochrane review to capture any studies published between September 2019 and November 2022. The search strategy used the search strategies set out in Appendix 2 of the aforementioned review (8). There were no language restrictions. We searched the following international electronic bibliographic databases: The Cochrane Library (Cochrane Central Register of Controlled Trials, Cochrane Database of Systematic Reviews), MEDLINE (Ovid, In-Process and Other Non-Indexed Citations Ovid, Epub ahead of print Ovid), EMBASE (Ovid), CINAHL Complete via EBSCOhost, Web of Science Social Sciences Citation Index, Science Citation Index, Conference Proceedings Citation Index-Science, Conference Proceedings Citation Index-Social Sciences), Epistemonikos (current issue), ClinicalTrials.gov, and WHO International Clinical Trials Registry Platform. In addition, we searched relevant regional databases. One of the authors (KRW) reviewed the titles and abstracts of all studies identified, to select all potentially relevant studies for full-text review. Full-text reports of all potentially relevant records were assessed against the inclusion and exclusion criteria detailed below.

We included prospective randomized controlled trials conducted in low- or middle-income countries, with MQ-LNS (∼250 kcal/d) provided to the intervention group for at least 3 mo between 6 and 23 mo of age. For a trial to be eligible for meta-analysis, a control group that did not receive MQ-LNS had to be available (e.g., comparator/control). The comparator group could have received other non-LNS type of child or household supplementation when the child was 6-23 mo of age, provided the intervention group received the same, e.g., household food ration, fortified blended food during the lean season, etc. Alternatively, the comparator group could have received SQ-LNS. Studies had to include longitudinal follow-up of each child, or repeated cross-sectional data collection. Studies were excluded if they focused primarily on the treatment, not prevention, of malnutrition, e.g., studies in which severe or moderate malnutrition was an inclusion criterion for children in the study. We also excluded studies conducted in a hospitalized population or among children with a pre-existing disease, and studies in which MQ-LNS provision was combined with additional supplemental food or nutrients for the child within a single arm (e.g., MQ-LNS + food rations vs. control), and there was no appropriate comparison group (e.g., food rations alone) that would allow separation of the MQ-LNS effect from effects of the other food or nutrients provided.

After initial screening, studies were grouped into 2 categories:

1. MQ-LNS used for < 6 mo (e.g., seasonal use during the lean season)
2. MQ-LNS used for ≥ 6 mo for children 6 - 23 mo of age

We conducted meta-analyses of studies in the second category, as the types and definitions of outcomes reported in studies in the first category were too heterogeneous for meta-analysis. We conducted two sets of meta-analyses. For the first, MQ-LNS arms were compared with control arms that received similar household-level or child-specific interventions. For the second meta-analysis, the comparator was arms that received SQ-LNS (<125 kcal/d).

### Outcomes assessed

The main outcomes for meta-analysis were weight-for-length Z (WLZ) score (sometimes measured as weight-for-height, WHZ), wasting (WLZ or WHZ < −2 SD), length-for-age Z (LAZ) score (sometimes measured as height-for-age, HAZ), and stunting (LAZ or HAZ < −2 SD). Additional outcomes considered (if available for more than 2 studies) were weight-for-age Z (WAZ) score, mid-upper arm circumference-for-age Z (MUACZ) score, head circumference-for-age Z score, cumulative incidence of wasting or low MUAC, underweight (WAZ < −2 SD), small head size (head circumference for age z-score < −2 SD), low MUAC (MUACZ < −2 SD or < 125 mm), acute malnutrition (WLZ < −2 SD or MUAC < 125 mm), severe wasting (WLZ < −3 SD), severe stunting (LAZ < −3 SD), very low MUAC (MUACZ < −3 SD or MUAC < 115 mm), and severe acute malnutrition (WLZ < −3 SD or MUAC < 115 mm). All z-scores were calculated using the 2006 WHO Child Growth Standards (17, 18). Each individual study’s principal endpoint was used for outcome variables.

### Data extraction

A standardized form was used to extract data from the publications of included studies for assessment of study quality and evidence synthesis. Two authors (CDA and KRW) extracted the data. Extracted information included: study setting, participant population and demographics (age at enrollment), study randomization scheme (cluster vs. individual), details of the intervention and control conditions (type and quantity of supplement, duration of supplementation, passive vs. active control, multi-component interventions, etc.), study sample size, primary and secondary outcomes, and information for the assessment of risk of bias.

### Risk of bias (quality) assessment

We assessed quality and risk of bias using the criteria outlined in the Cochrane Handbook for Systematic Reviews of Interventions (19) and the Grading of Recommendations Assessment, Development and Evaluation (GRADE) framework (20). Two reviewers (CDA and KRW) independently assessed the risk of bias using the following criteria: random sequence generation, allocation concealment, blinding of participants and personnel, blinding of outcome assessment, incomplete outcome data, selective reporting and other sources of bias. GRADE criteria included: risk of bias, inconsistency of effect, imprecision, indirectness, and publication bias.

### Data synthesis and analysis

We used R version 4.1.1 (R Core Team, Vienna, Austria) for all statistical analyses. For each study, intervention groups were combined into LNS and Control groupings by taking the weighted average for means and standard deviations and by using the summed event count and total denominator sample size for binary outcomes. We extracted the endline prevalences and event counts from study publications and completed analysis based on the extracted values. If event count was not available for binary outcomes, it was approximated based on reported prevalence. If standard deviation values were not available for continuous outcomes for a given study, they were approximated based on the weighted averages of standard deviations for the same outcomes within intervention groups reported from comparable trials. Standardized mean difference-in-differences (MD) (from time of start of child supplementation) were estimated and pooled for continuous outcomes, and prevalence ratios (PR) and prevalence differences (PD) were estimated and pooled for binary outcomes, recalibrating to account for baseline differences as needed. Pooling was done using a random effect inverse variance weighting approach and confidence intervals were two-sided at the 95% confidence level. All testing was two-sided and considered significant at the α=0.05 level, unless otherwise specified.

We explored trial protocol heterogeneity by summarizing the methods, participants, interventions, monitoring approaches, and potential for bias of each study. Studies with protocols that differed substantially from the rest were assessed to determine if they differed in treatment effect, via sensitivity analyses. For statistical heterogeneity we calculated I² statistics.

### Estimated nutrient intakes from MQ-LNS and SQ-LNS under different scenarios

To understand the implications of incomplete consumption of the daily ration of MQ-LNS, we calculated the amounts of each nutrient that would be consumed if children ate the full 50g sachet or only half of the sachet (i.e., 25 g/d) of the MQ-LNS formulation used by WFP. For comparison, we also calculated nutrient intakes based on consuming a full sachet of SQ-LNS (20 g/d). **Supplemental Table 1** shows the nutrient content of the WFP formulations of MQ-LNS and SQ-LNS, as well as the SQ-LNS formulation used in the most recent research trials (which differs slightly from the WFP formulation), along with the recommended nutrient intake (RNI) values used for these calculations.

## Results

### Literature search and trial characteristics

We screened 2,709 records and assessed 58 reports that met initial screening criteria (**Figure 1**). Of those 58, 24 were excluded because they were not randomized trials, did not use MQLLNS, were focused only on treating malnutrition, had no appropriate comparison group, were ongoing trials or were systematic reviews. The remaining 34 reports were publications from 11 different studies. Of those 11 studies, 3 were categorized as short-term use of MQ-LNS (< 6 mo) and 8 were categorized as using MQ-LNS for ≥ 6 mo.

**Figure 1:**
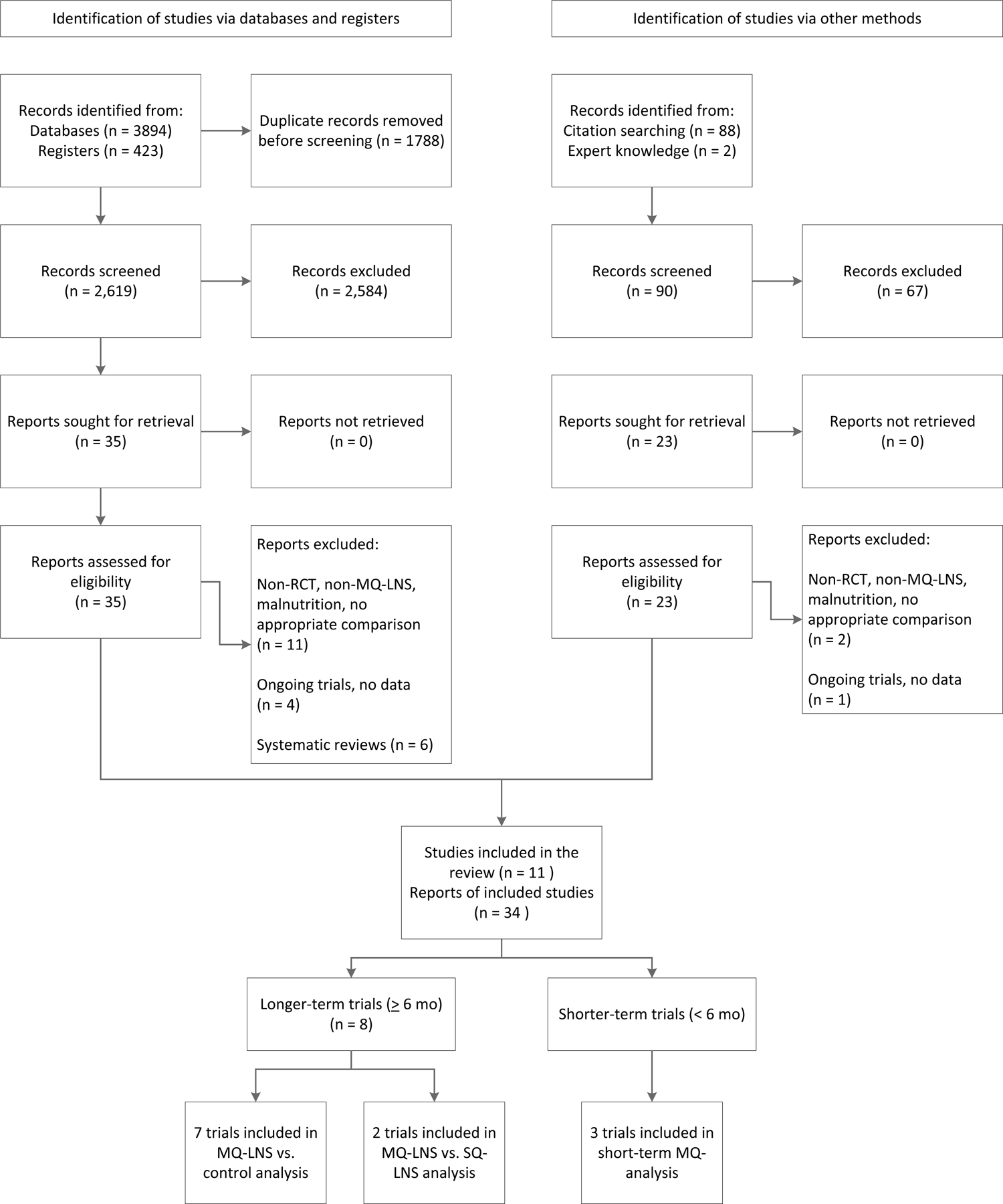
Study flow diagram. MQ-LNS, medium-quantity lipid-based nutrient supplements, SQ-LNS, small-quantity lipid-based nutrient supplements; RCT, randomized controlled trial.

**Table 1** shows the characteristics of the trials in these two categories. The 3 short-term trials were conducted in Chad, Niger and Nigeria (21–23), and all were designed to provide MQ-LNS only during the lean season (generally for 3-5 mo) to children who ranged in age from 6 to 24 mo (2 trials) (22, 23) or 6 to 36 mo (1 trial) (21). All 3 were cluster-randomized trials, but the number and types of intervention groups varied. In Chad there were 2 groups: all households received a monthly food package and the intervention households received an additional ration of daily MQ-LNS for the child aged 6-36 mo. In Niger, there were 7 groups in the full study but 5 of them received LQ-LNS or Supercereal+ as the child’s food ration, leaving 2 groups for the comparisons herein: MQ-LNS + cash transfer vs. cash transfer only. In Nigeria, there were 2 groups: all children received seasonal malaria chemoprevention and the intervention group also received MQ-LNS. The same MQ-LNS formulation (46 g) was used in all 3 trials, but in Nigeria the daily amount was a half-ration (23 g) for infants 6-11 mo of age and a full ration for children 12-24 mo of age. Two trials (Chad and Nigeria) presented results relevant to both wasting and linear growth, but the Niger study provided data only on acute malnutrition. Because the definition of wasting was different across trials, there were no comparisons for which we had > 2 studies, and thus we did not conduct a meta-analysis of the short-term trial results (the findings are presented in narrative form below).

**Table 1.** Characteristics of studies included in the systematic review and meta-analysis

| Country, years of study, study name, author date | Study design | Intervention groups<sup>1</sup> | LNS product(s) | Age at start of supplementation (duration of supplementation) | Outcomes assessed |
| --- | --- | --- | --- | --- | --- |
| **MQ-LNS used for < 6 mo** |  |  |  |  |  |
| Chad, 2010, (Huybregts 2012) (21) | Cluster randomized controlled pragmatic trial, longitudinal follow-up | **(1) Intervention (n=458) vs. (2) control (n=613):** All households received a monthly food package. Intervention households (child 6-36 mo) also received MQ-LNS for 4 mo (June - October). | MQ-LNS (Plumpy'Doz, 46 g, 250 kcal/d) | 6-36 mo (4 mo) | Change in WHZ, HAZ, MUAC; prevalence of stunting; cumulative incidence of acute malnutrition or wasting |
| Niger, 2011, (Langendorf 2014) (22) | Cluster partially-randomized controlled trial, longitudinal follow-up | (1) LQ-LNS, (2) LQ-LNS + cash transfer, **(3) MQ-LNS + cash transfer n=1089**, (4) Supercereal plus + cash transfer, (5) Supercereal plus + family food ration, (6) Supercereal plus, **(7) cash transfer only (n=680)**. LNS distributed August – December. | MQ-LNS (Plumpy'Doz, 46 g, 250 kcal/d); LQ-LNS (Supplementary Plumpy, 92 g, 500 kcal/d) | ~6-24 mo (5 mo max) | Incidence of SAM and MAM |
| Nigeria, 2014, (Ward 2019) (23) | Two-cluster randomized controlled pragmatic trial, repeated cross-sectional household surveys | **(1) Seasonal malaria chemoprevention (SMC) (n=803) vs. (2) SMC + SQ/MQ-LNS (n=650).** LNS distributed August – November. | MQ-LNS (Plumpy'Doz, half-dose from 6-11 mo (~23 g, 125 kcal/d); full-dose 12-24 mo (46 g, 250 kcal/d)) | 6-24 mo (4 mo) | WLZ, LAZ, WAZ, MUACZ; prevalence of wasting, stunting, underweight, low MUAC and SAM |
| **MQ-LNS used for > 6 mo for children 6 - 23 mo of age** |  |  |  |  |  |
| Bangladesh, 2012-2014, JiVitA-4, (Christian 2015) (24) | Cluster RCT, longitudinal follow-up | **(1) SQ/MQ-LNS (n=3160), (2) control (n=1438),** (3) wheat-soy blend. All groups received IYCF counseling. | MQ-LNS (Plumpy'Doz, chickpea-based LNS or rice-lentil LNS): 1/2 dose from 6-11 mo (~23 g, 125 kcal/d), full-dose 12-18 mo (46 g, 250 kcal/d)) | 6 mo (12 mo) | Change in WLZ, LAZ, , WAZ; prevalence of wasting, stunting, underweight; cumulative incidence of wasting |
| Malawi, 2004- | RCT, longitudinal | **(1) SQ-LNS (n=60), (2) MQ-LNS** | SQ-LNS (25 g, 130 kcal/d) | 6 mo (12 mo) | Change in WLZ, LAZ, WAZ; |
| 2005, (Phuka 2008) (25) | follow-up | (n=61), (3) micronutrient fortified maize-soy flour | and MQ-LNS (50 g, 260 kcal/d); peanut- and milk-based LNS; same amounts of micronutrients (except Ca) provided to each group |  | incidence of severe or moderate to severe wasting, stunting and underweight |
| Malawi, 2008-2010, (Mangani 2015) (26) | RCT, longitudinal follow-up | **(1) MQ-LNS containing milk (n=212), (2) MQ-LNS without milk (n=210), (3) corn-soy blend, (4) control (delayed intervention, n=209))** | MQ-LNS (54 g, 280 kcal/d); peanut + milk or soy based LNS | 6 mo (12 mo) | Change in WLZ, LAZ, WAZ; incidence of severe wasting, any stunting, severe stunting, very severe stunting (LAZ < -3.5 SD), severe underweight;; prevalence of severe wasting, any stunting, severe stunting, and very severe stunting |
| Malawi, 2009-2012, (Maleta 2015) (13) | RCT, longitudinal follow-up | **(1) SQ-LNS containing milk (10 g/d, n=321), (2) SQ-LNS containing milk (20 g/d, n=322), (3) SQ-LNS without milk (20 g/d, n=323), (4) MQ-LNS containing milk (40 g/d, n=322), (5) MQ-LNS without milk (40 g/d, n=324), (6) control (n=320)** | SQ-LNS with or without milk (10 or 20 g/d, ~55-118 kcal/d) and MQ-LNS with or without milk (40 g, 240 kcal/d); same amounts of micronutrients provided to each group | 6 mo (12 mo) | Change in WLZ, LAZ, WAZ, MUACZ; incidence of wasting, severe wasting; stunting, severe stunting; underweight, severe underweight |
| Mali, 2013-2016, (Adubra 2019) (27, 28) | Cluster RCT, repeated cross-sectional surveys | **(1) Community health & nutrition program (SNACK) (n=1278), (2) SNACK + Cash n=1223), (3) SNACK + MQ-LNS (n=1289), (4) SNACK + Cash + MQ-LNS (n=1256)** | MQ-LNS (Plumpy'Doz, 46 g, 250 kcal/d) | 6 mo (18 mo) | WLZ, HAZ ; prevalence of wasting, stunting |
| Pakistan (Sindh Cohort 2), 2014-2016, (Khan 2020) (29) | Cluster RCT, longitudinal follow-up | **(1) MQ-LNS (n=419) vs. (2) standard of care (n=451)** | MQ-LNS (Wawamum, 50 g, 255 kcal/d); chickpea- and milk-based formulation | 6-18 mo (6-18 mo) | Prevalence of wasting, stunting, underweight |
| Pakistan (Sindh Cohort 1), 2015-2018, (Soofi 2022) (30) | Cluster RCT, longitudinal follow-up | **(1) WSB+ in pregnancy and 6 mo post-partum, child MQ-LNS 6-23 mo (n=705) vs. (2) standard of care (n=753)** | MQ-LNS (Wawamum, 50 g, 255 kcal/d); chickpea- and milk-based formulation | 6 mo (18 mo) | WLZ, LAZ, WAZ; prevalence of wasting, stunting, underweight |
| Pakistan (Punjab), 2017-2019, (Soofi | Cluster RCT, longitudinal follow-up | **(1) Unconditional cash transfer (UCT, n=434), (2) UCT + SBCC (n=433), (3) UCT + MQ-LNS** | MQ-LNS (Wawamum, 50 g, 225 kcal/d); chickpea- and milk-based formulation | 6 mo (18 mo) | WLZ, LAZ, WAZ; prevalence of wasting, stunting, underweight |
| 2022) (31) |  | **(n=430), (4) UCT + MQ-LNS + SBCC (n=432)** |  |  |  |

HAZ, height-for-age z-score; IYCF, infant and young child feeding; LAZ, length-for-age z-score; LNS, lipid-based nutrient supplement; LQ-LNS, large-quantity lipid-based nutrient supplement; MAM, moderate acute malnutrition; MQ-LNS, medium-quantity lipid-based nutrient supplement; MUAC, mid-upper arm circumference; MUACZ, mid-upper arm circumference z-score; RCT, randomized controlled trial; SAM, severe acute malnutrition; SBCC, social and behavior change communication; SMC, seasonal malaria chemoprevention; SQ-LNS, small-quantity lipid-based nutrient supplement; UCT, unconditional cash transfer; WAZ, weight-for-age z-score; WHZ, weight-for-height z-score; WLZ, weight-for-length z-score; WSB+, wheat-soy blend plus.
<sup>1</sup>Intervention groups used in this meta-analysis are in bold; sample sizes (n) are the number of children randomized to each group, except for the study in Nigeria (Ward) for which the numbers of children in the midline survey are listed.

The 8 trials that used MQ-LNS for ≥ 6 mo were conducted in Bangladesh (1 trial), Malawi (3 trials), Mali (1 trial) and Pakistan (3 trials) (13, 24–31). In most of the trials, children began receiving MQ-LNS at 6 mo of age and the duration of supplementation was 12-18 mo (i.e., until 18-24 mo of age); the exception was one study in Pakistan (29) in which children varied in age at enrollment (6-18 mo) and received MQ-LNS for 6-18 mo, i.e., until 24 mo of age. The 3 studies in Malawi were randomized controlled trials with longitudinal follow-up, and the other 5 studies were cluster-randomized trials (4 with longitudinal follow-up and 1 with repeated cross-sectional surveys). All trials included at least one intervention group in which children received MQ-LNS (40-54 g per ration), but in the Bangladesh trial (24), the daily amount was a half-ration (23 g) for infants 6-11 mo of age and a full ration for children 12-24 mo of age. In 6 of the 8 trials, there was at least one control group in which children did not receive any food supplement; these control groups received standard of care, infant and young child feeding (IYCF) counseling and/or delayed intervention except for 1 trial (in Punjab, Pakistan) (31) in which all households (both control and intervention groups) received a cash transfer. The other 2 trials included a study in Malawi (25) in which the comparison groups received either SQ-LNS or a fortified maize-soy flour, and the study in Mali (27) in which all households received a package of interventions that included blanket provision of a fortified blended flour during the lean season for children 6-23 mo of age. In 1 trial in Pakistan (30), the intervention included maternal food supplementation (with wheat-soy blend) in addition to the MQ-LNS provided to children. Prevalence of wasting in the control groups at endline was relatively high: 8% in Mali (27, 28), 9% in Malawi (13), 16% in Bangladesh (24), and 10% (31), 21% (29) and 28% (30) in Pakistan.

We evaluated risk of bias in the 8 longer-duration trials that were included in the meta-analysis (**Supplemental Table 2**, **Supplemental Figure 1**). All trials were judged to have low risk of bias for random sequence generation, allocation concealment, incomplete outcome, selective reporting, and “other.” All trials were judged to have high risk of bias for blinding of participants, because blinding was not possible given the nature of the intervention. Risk of bias in outcome assessment was mixed (3 low, 5 high).

### Anthropometric outcomes

#### Short-term trials providing MQ-LNS during the lean season

In Chad (21), where all households were included in a general food distribution program, there were no significant differences between the MQ-LNS and control groups in mean WHZ (SD) [−1.05 (0.93) vs. −1.09 (0.95), p=0.89], change in MUAC (+0.01 Z-score/mo; 95% CI: −0.02, 0.04; p = 0.49) or incidence of wasting [incidence rate ratio (95% confidence interval (CI)): 0.86 (0.67, 1.11), p=0.25]. The intervention group had a greater gain in HAZ (+0.03 Z-score/mo; 95% CI: 0.01, 0.04; p<0.001), but stunting prevalence at endline did not differ significantly between groups (46.2 vs 52.3%, odds ratio (95% CI): 0.69 (0.45, 1.07), p=0.099).

In Niger (22), the only outcomes reported were moderate acute malnutrition (MAM, defined as − 3 ≤ WLZ < −2 and/or 11.5 ≤ MUAC < 12.5 cm) and severe acute malnutrition (SAM, defined as WLZ < −3 and/or MUAC < 11.5 cm and/or bipedal edema). Compared to cash transfer alone, provision of MQ-LNS reduced the risk of both MAM and SAM [Cash vs. MQ-LNS+cash, MAM Hazard Ratio = 2.07 (95% CI: 1.52, 2.82), SAM Hazard Ratio = 2.12 (95% CI: 1.26, 3.58)].

In Nigeria (23), there was no significant effect of MQ-LNS on mean change (95% CI) in WLZ [−0.41 (−1.01, 0.20)], LAZ [0.16 (−0.44, 0.76)], WAZ [−0.08 (−0.57, 0.42)] or MUACZ [0.26 (−0.12, 0.64)], or on the prevalence [PR (95% CI)] of wasting [1.35 (0.69, 2.62), stunting [0.94 (0.45, 1.97), underweight [0.97 (0.52, 1.80)] or low MUAC [1.93 (0.17, 22.41)] between the baseline and midline surveys (i.e., at the end of the 4-mo MQ-LNS distribution period). Between baseline and endline surveys (> 6 mo after MQ-LNS distribution had ended), there were also no significant differences in growth outcomes between groups except that the MQ-LNS group had a significantly greater change in MUACZ [0.60 (0.26, 0.94), p<0.01].

#### Longer-term trials providing MQ-LNS for ≥ 6 mo

The outcomes available for > 2 of the 8 longer-term trials, and thus included in the meta-analysis, were WLZ, wasting, LAZ, stunting, WAZ, and underweight. The comparisons of MQ-LNS vs. Control are shown in **Table 2**. There were significant effects of MQ-LNS on WLZ [MD +0.09 (95% CI: 0.05, 0.13)], wasting [PR 0.89 (95% CI: 0.81, 0.97), PD −1.4 (−2.4, −0.4)] and underweight [PR 0.94 (0.88, 0.99)], but not on LAZ, stunting or WAZ. **Figures 2 and 3** show the forest plots for wasting and stunting PRs, respectively, for MQ-LNS vs Control, and **Supplemental Figures 2-5** show the forest plots for WLZ, LAZ, WAZ and underweight PR. We rated the quality of the evidence for all outcomes as moderate based on GRADE criteria: risk of bias was generally low, heterogeneity was generally low to moderate (Table 2), all trials were directly aimed at evaluating MQ-LNS, and funnel plots revealed no indication of publication bias, but there were relatively few trials (4-8 depending on the outcome) and only 4 countries were represented.

**Figure 2:**
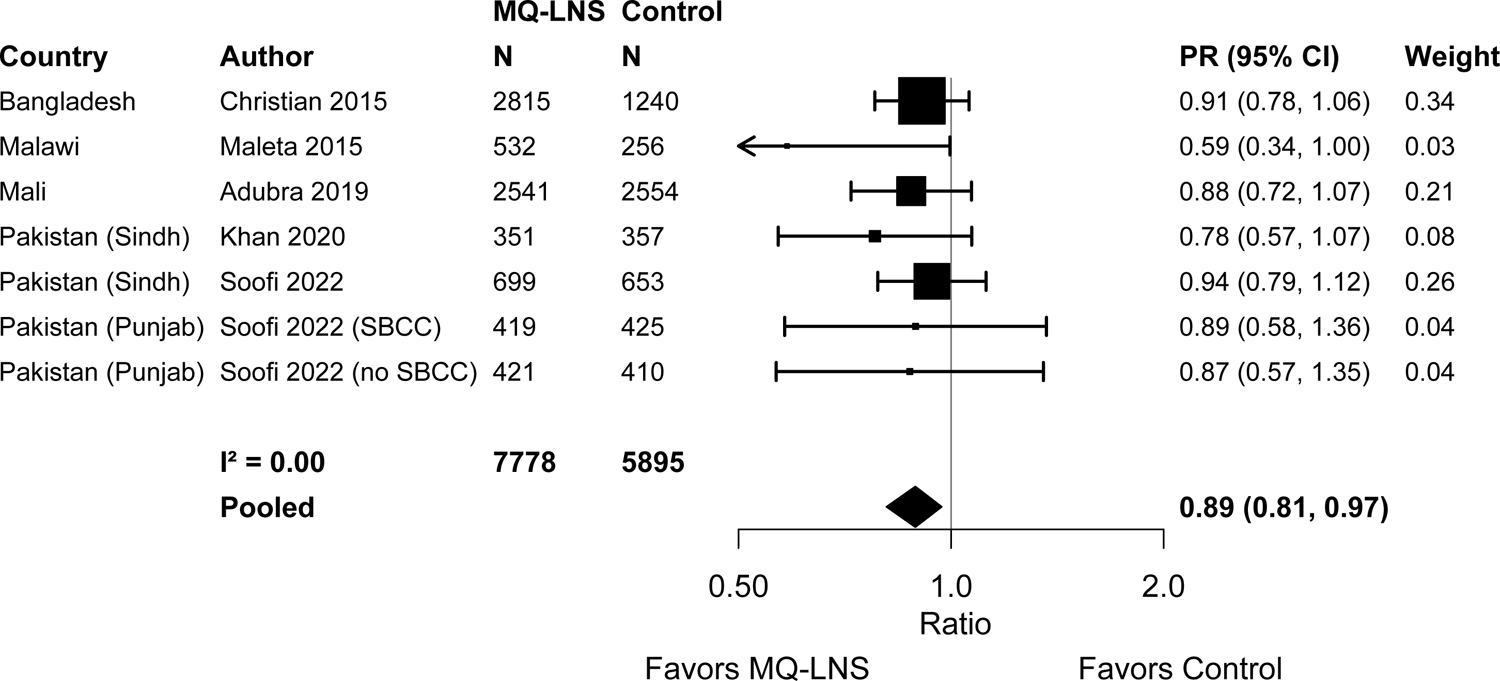
Forest plot of effect of MQ-LNS vs. control on wasting prevalence. MQ-LNS, medium-quantity lipid-based nutrient supplements; PR, prevalence ratio. Individual study prevalence estimates were extracted from published material and prevalence ratios were pooled using inverse-variance weighting with random effects. For Adubra (27, 28), values are based on a combined weighted average of the mean change for the SNACK+LNS and SNACK + CASH + LNS groups vs the combined weighted average of the mean change for the SNACK and SNACK + CASH groups. For Soofi (Sindh) (30), PRs were calculated after recalibrating the endline prevalence differences to take into account the intervention group difference already present at 6 mo, so the effect estimates are related only to the effects of child supplementation.

**Figure 3:**
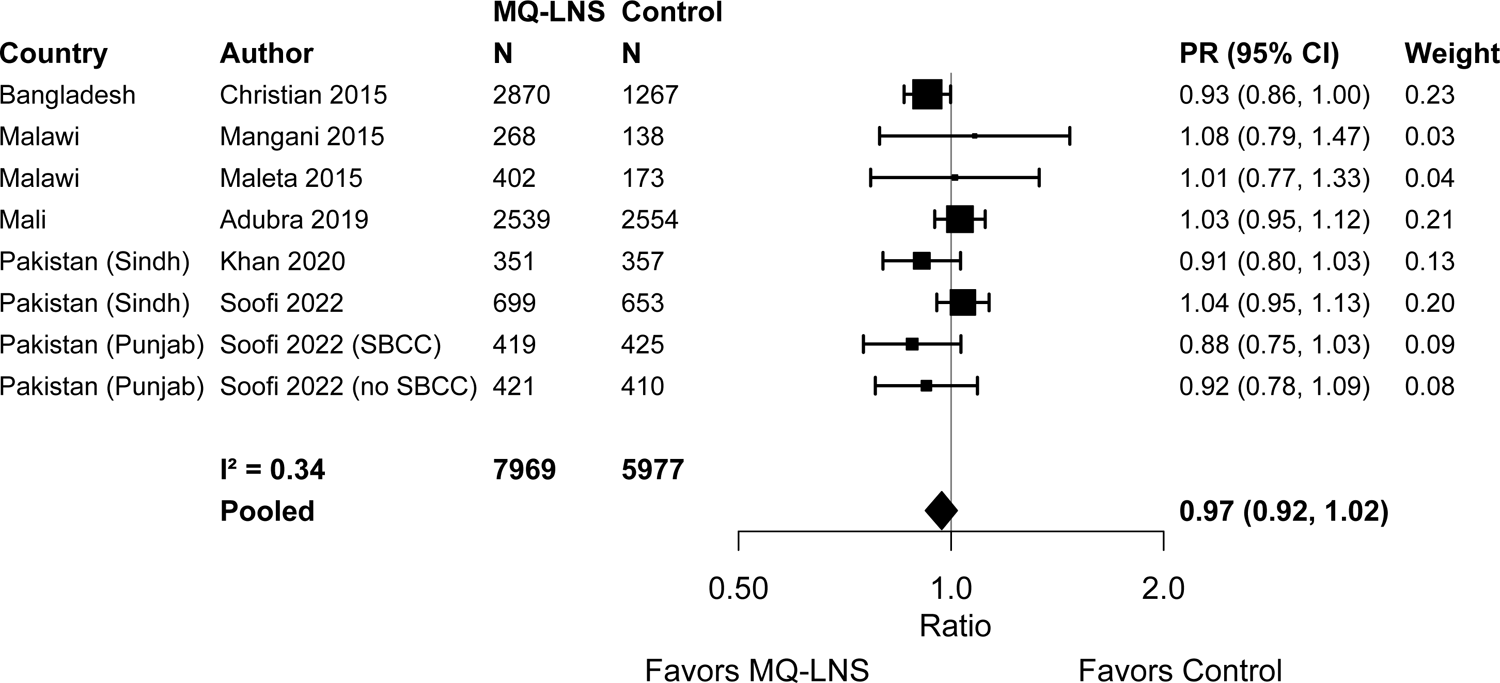
Forest plot of effect of MQ-LNS vs. control on stunting prevalence. MQ-LNS, medium-quantity lipid-based nutrient supplements; PR, prevalence ratio. Individual study prevalence estimates were extracted from published material and prevalence ratios were pooled using inverse-variance weighting with random effects. For Soofi (Sindh) (30), PRs were calculated after recalibrating the endline prevalence differences to take into account the intervention group difference already present at 6 mo, so the effect estimates are related only to the effects of child supplementation.

**Table 2.** Main effects of MQ-LNS (relative to Control) and SQ-LNS (relative to Control) on growth outcomes^1^

| Outcome | MQ-LNS vs Control |  |  | SQ-LNS vs Control<br>(from IPD meta-analysis (32)) |  |  |
| --- | --- | --- | --- | --- | --- | --- |
|  | N (# comparisons)<sup>2</sup> | MD, PR or PD<sup>3</sup> (95% CI) | *I*<sup>2(4)</sup> | N (# comparisons) | MD, PR or PD<sup>3</sup> (95% CI) | *I*<sup>2(4)</sup> |
| WLZ (MD) | 11,603 (5) | **0.09**<br>(0.05, 0.13)<br>P<0.001 | 0.03 | 36,608 (17) | **0.08**<br>(0.06, 0.10)<br>P<0.001 | 0.51 |
| Wasting (PR) | 13,673 (7) | **0.89**<br>(0.81, 0.97)<br>P=0.008 | 0.00 | 36,311 (16) | **0.86**<br>(0.80, 0.93)<br>P<0.001 | 0.00 |
| Wasting (PD) | 13,673 (7) | **-1.4**<br>(-2.4, -0.4)<br>P=0.008 | 0.00 | 36,311 (16) | **-0.6**<br>(-1.0, -0.1)<br>P=0.01 | 0.12 |
| LAZ (MD) | 11,812 (5) | 0.04<br>(-0.02, 0.11) | 0.56 | 36,795 (17) | **0.14**<br>(0.11, 0.16)<br>P<0.001 | 0.65 |
| Stunting (PR) | 13,954 (8) | 0.97<br>(0.92, 1.02) | 0.34 | 36,795 (17) | **0.88**<br>(0.85, 0.91)<br>P<0.001 | 0.49 |
| Stunting (PD) | 13,954 (8) | -1.2<br>(-3.5, 1.0) | 0.32 | 36,795 (17) | **-5.0**<br>(-5.9, -4.1)<br>P<0.001 | 0.54 |
| WAZ (MD) | 6,683 (4) | 0.07<br>(-0.03, 0.17) | 0.63 | 36,787 (17) | **0.13**<br>(0.11, 0.15)<br>P<0.001 | 0.66 |
| Underweight (PR) | 8,550 (6) | **0.94**<br>(0.88, 0.99)<br>P=0.028 | 0.06 | 36,787 (17) | **0.87**<br>(0.83, 0.97)<br>P<0.001 | 0.42 |
| Underweight (PD) | 8,550 (6) | -0.7<br>(-4.2, 2.7) | 0.66 | 36,787 (17) | **-3.1**<br>(-3.8, -2.3)<br>P<0.001 | 0.60 |

<sup>1</sup>LAZ, length-for-age z score; MQ-LNS, medium-quantity lipid-based nutrient supplement; MD, mean difference; PD, prevalence difference (percentage points); PR, prevalence ratio; WAZ, weight-for-age z score; WLZ, weight-for-length z score.
<sup>2</sup>The maximum number of comparisons was 8 (7 studies, with 2 comparisons for 1 of those studies (31)), for stunting. For the other outcomes, prevalence of wasting was not reported in 1 study (26), prevalence of underweight was not reported in 2 studies (26, 27), and z-score values from some studies were not available (29) or were not calculable as difference-in-difference estimates (31).
<sup>3</sup>For continuous outcomes, values are MDs: MQ-LNS – control (95% CIs). For binary outcomes, values are PR and PDs: MQ-LNS compared with control (95% CIs). P-values correspond to the pooled main effect 2-sided superiority testing of the intervention effect estimate and 95% CI.
<sup>4</sup>*I*<sup>2</sup> describes the percentage of variability in effect estimates that may be due to heterogeneity rather than chance. Roughly, 0.3–0.6 may be considered moderate heterogeneity.

Because the trial in Bangladesh used a half-ration of MQ-LNS for children 6-11 mo of age, we conducted a sensitivity analysis excluding that trial, and results remained significant for WLZ [0.08 (0.02, 0.14)], and wasting [PR 0.87 (0.78, 0.98), PD −1.4 (−2.5, −0.2)], became non-significant for underweight PR [0.97 (0.90, 1.04)], and remained non-significant for LAZ [0.01 (−0.04, 0.05)], stunting [PR 0.98 (0.93, 1.04)] and WAZ [0.01 (−0.07, 0.09)]. We also conducted a sensitivity analysis excluding the trial in Pakistan in which both mothers and children received supplementation (30), and the results were similar for WLZ [0.10 (95% CI: 0.06, 0.14)], wasting [PR 0.87 (0.78, 0.96), PD −1.4 (−2.5, −0.3)] and underweight [PR 0.93 (0.86, 1.01)], and remained non-significant for LAZ [0.05 (−0.02, 0.13)], stunting [PR 0.95 (0.90, 1.01)] and WAZ [0.10 (−0.02, 0.21)].

For comparison, Table 2 also shows the effects of SQ-LNS on the same outcomes, from the previously reported meta-analyses (32). Effects of MQ-LNS on WLZ and wasting PR were similar to those of SQ-LNS; for example, the relative reduction in wasting prevalence was 11% for MQ-LNS and 14% for SQ-LNS. There was a slightly greater PD for MQ-LNS (−1.4 percentage points) than for SQ-LNS (−0.6), despite a similar PR, because the wasting prevalence was generally higher in the MQ-LNS study sites (8-28%) than in the SQ-LNS sites (1-15%) (32). For LAZ, WAZ and stunting, however, significant effects were demonstrated for SQ-LNS but not for MQ-LNS. For underweight, effects of MQ-LNS (6% relative reduction) were significant but of smaller magnitude than those for SQ-LNS (13% relative reduction), and there was no significant effect of MQ-LNS on the PD for underweight.

Two of the 8 trials (25, 26) allowed for direct comparisons of SQ-LNS and MQ-LNS groups, shown in **Table 3**. There were no significant differences in any of these growth outcomes between groups. A sensitivity analysis was conducted to restrict the type of LNS to milk-containing formulations, which showed similar results (Table 3). For this comparison, we rated the quality of the evidence for all outcomes as low because there were only 2 trials, both conducted in Malawi.

**Table 3.**
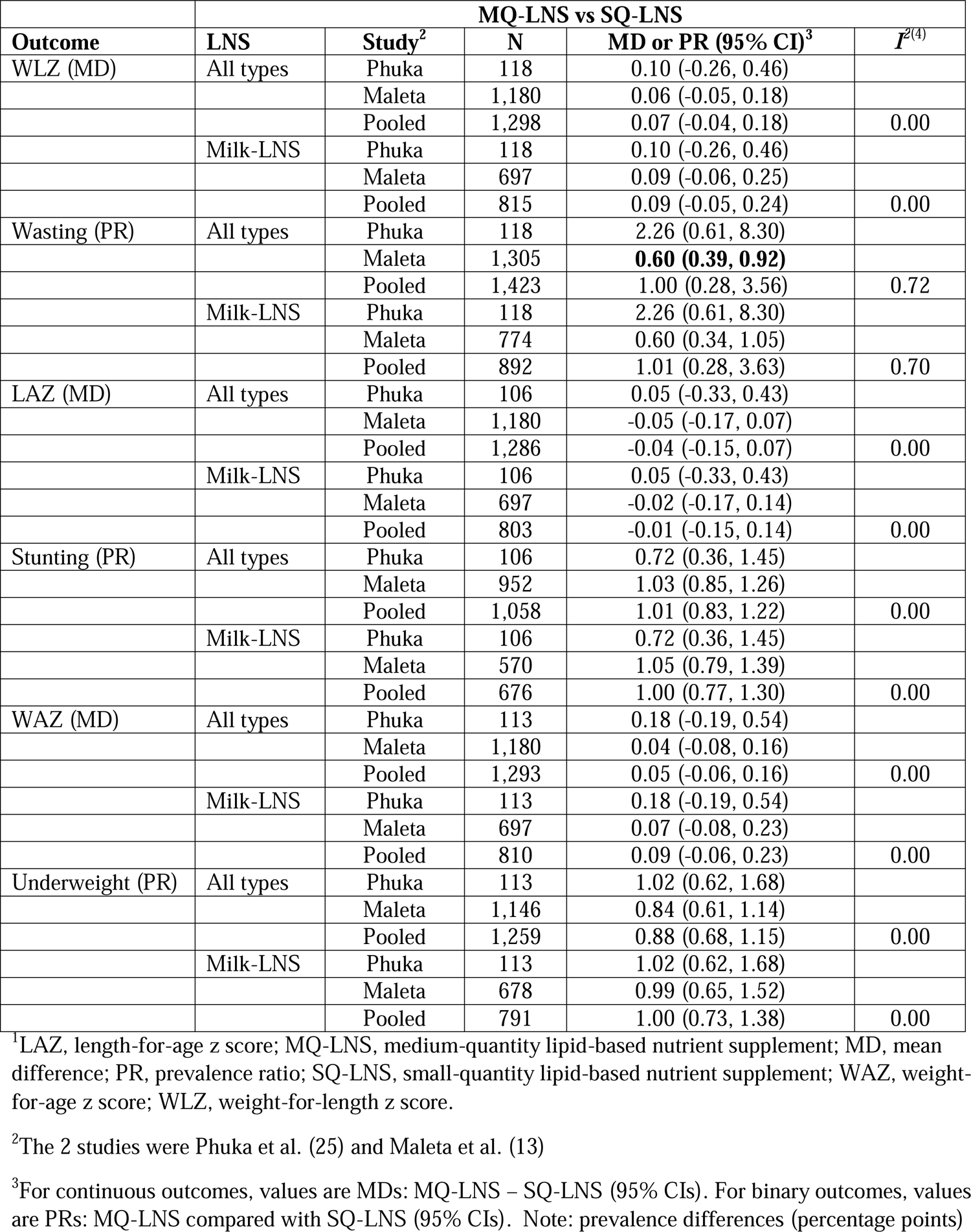
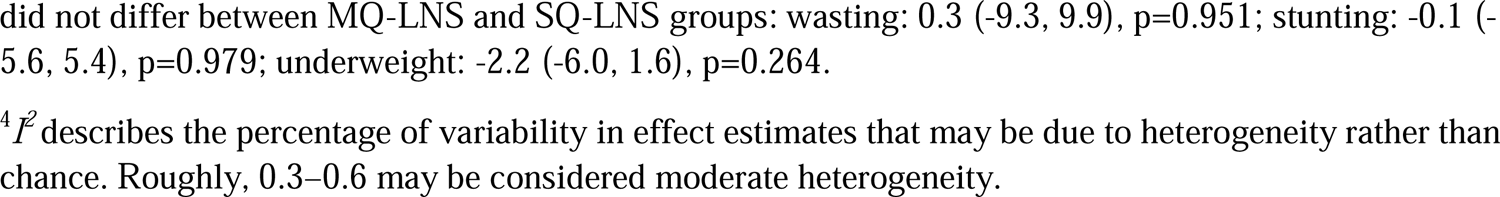
Effects of MQ-LNS vs SQ-LNS on growth outcomes in trials with direct comparisons of these 2 interventions**^1^**

#### Estimated nutrient intakes from MQ-LNS under different scenarios

Supplemental Table 1 shows the estimated nutrient intakes, as a percentage of the RNI, from a half-ration of MQ-LNS (25 g/d) or a full ration of SQ-LNS (20 g/d). In terms of nutrient shortfalls, the most problematic nutrients (**Table 4**) are iron and calcium: the half-ration of MQ-LNS would provide only 23% (at 6-12 mo) and 36% (at 12-23 mo) of the RNI for iron, and only 48% (at 6-12 mo) and 30% (at 12-23 mo) of the RNI for calcium. The half-ration of MQ-LNS would also fall short of the RNI for zinc (64% of the RNI at 12-23 mo), vitamin A (70% of the RNI at both 6-12 and 12-23 mo), and thiamin (83% of the RNI at 6-12 mo and 50% of the RNI at 12-23 mo).

**Table 4.**
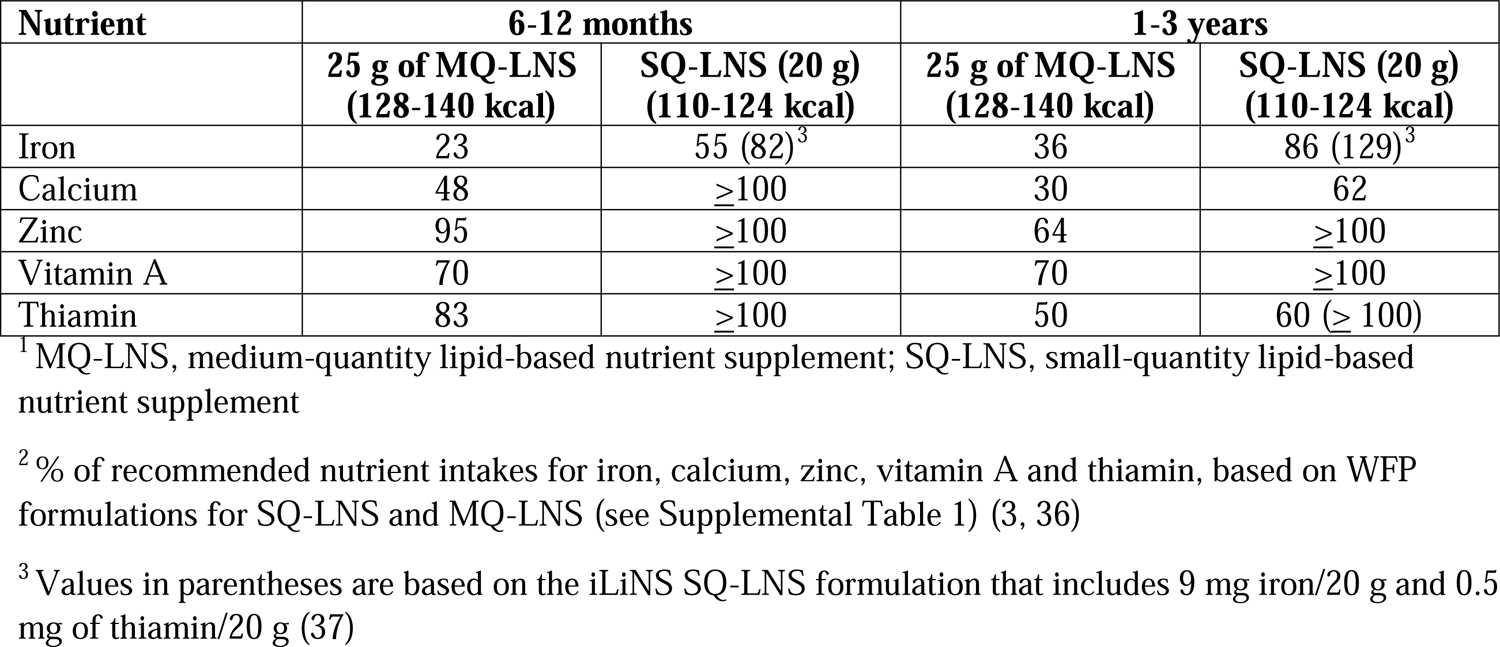
Percentage of recommended nutrient intakes for several key nutrients provided by a half-ration of MQ-LNS or a full ration of SQ-LNS^1,2^

| Nutrient | 6-12 months |  | 1-3 years |  |
| --- | --- | --- | --- | --- |
|  | 25 g of MQ-LNS<br>(128-140 kcal) | SQ-LNS (20 g)<br>(110-124 kcal) | 25 g of MQ-LNS<br>(128-140 kcal) | SQ-LNS (20 g)<br>(110-124 kcal) |
| Iron | 23 | 55 (82)<sup>3</sup> | 36 | 86 (129)<sup>3</sup> |
| Calcium | 48 | ≥100 | 30 | 62 |
| Zinc | 95 | ≥100 | 64 | ≥100 |
| Vitamin A | 70 | ≥100 | 70 | ≥100 |
| Thiamin | 83 | ≥100 | 50 | 60 (≥ 100) |

<sup>1</sup> MQ-LNS, medium-quantity lipid-based nutrient supplement; SQ-LNS, small-quantity lipid-based nutrient supplement
<sup>2</sup> % of recommended nutrient intakes for iron, calcium, zinc, vitamin A and thiamin, based on WFP formulations for SQ-LNS and MQ-LNS (see Supplemental Table 1) (3, 36)
<sup>3</sup> Values in parentheses are based on the iLiNS SQ-LNS formulation that includes 9 mg iron/20 g and 0.5 mg of thiamin/20 g (37)

## Discussion

In this systematic review of studies in which MQ-LNS was used for prevention of child undernutrition, we identified 3 eligible trials that examined supplementation for 3-5 mo during the lean season and were targeted at children 6-24 or 6-36 mo of age, and 8 longer-duration trials that generally provided MQ-LNS starting at 6 mo of age for a duration of 6-18 mo and met our criteria for meta-analysis. The 3 short-term trials did not show consistent effects on child growth outcomes. In the meta-analysis of the longer-term trials, there was a positive effect on WLZ (+0.09) and relative reductions in wasting (11%) and underweight (6%) in the MQ-LNS groups compared to control, but no effects on LAZ or stunting. Previous meta-analyses of SQ-LNS vs control also showed effects on WLZ (+0.08), wasting (14% reduction) and underweight (13% reduction), but SQ-LNS also had an impact on LAZ (+0.14) and stunting (12% reduction) (32). In two trials that directly compared SQ-LNS and MQ-LNS, there were no significant differences in these growth outcomes. These results suggest no evidence of superiority of MQ-LNS over SQ-LNS for prevention of wasting, and less impact of MQ-LNS on linear growth and stunting.

The evidence base for MQ-LNS is considerably smaller and more heterogeneous than is the case for SQ-LNS. The MQ-LNS vs. control meta-analysis results are based on 4-7 trials (depending on the outcome) in 4 countries and a total sample size of <14,000, whereas the SQ-LNS vs. control meta-analysis is based on 14 trials in 9 countries and a total sample size of ∼37,000 children. The evidence base for short-term MQ-LNS trials aimed at prevention of seasonal undernutrition is very small, with only 3 trials and a total of only ∼4,300 children. Results of those 3 trials were mixed. In Niger, the MQ-LNS group had a lower risk of both MAM and SAM, but in Chad and Nigeria, there were no significant effects on WLZ/WHZ or wasting. There were positive effects of MQ-LNS on linear growth in Chad, but no such effects were observed in Nigeria and linear growth outcomes were not reported in Niger. Thus, it is difficult to draw conclusions about the effectiveness of short-term seasonal supplementation for prevention of child undernutrition.

The evidence for direct comparisons of SQ-LNS and MQ-LNS is limited to 2 trials, both conducted in Malawi in contexts in which there has been little impact of either supplement (compared to control groups) on growth outcomes (25, 26). The reasons for this lack of response in Malawi are unclear but may be related to high levels of inflammation and infection among infants and young children in this setting (33), which may constrain a growth response to nutritional supplementation. Additional studies directly comparing SQ-LNS and MQ-LNS for prevention of child undernutrition in other settings are needed.

The contexts in which the MQ-LNS and SQ-LNS trials were conducted varied considerably. Most of the MQ-LNS trials were conducted in countries with high levels of food insecurity. Among the 14 SQ-LNS trials, 7 were conducted in populations in which >30% of households reported moderate-to-severe food insecurity at baseline (32). Although the level of food insecurity could potentially influence the effectiveness of supplementation, the IPD meta-analysis indicated that household food insecurity did not modify the effects of SQ-LNS on WLZ, LAZ, wasting or stunting (32). In other words, the impact of SQ-LNS on these outcomes was evident even among children in households with moderate-to-severe food insecurity. Thus, it seems unlikely that greater levels of food insecurity in the MQ-LNS trials explains their lack of impact (vs. control) on linear growth or stunting.

Four of the 8 longer-term MQ-LNS trials were carried out within community-based programs that included other components, such as health and nutrition services (27, 29, 30), cash transfers (31) or maternal nutritional supplementation (30). Similarly, 6 of the 14 SQ-LNS trials were conducted within existing community-based or clinic-based programs. Therefore, the meta-analysis results for both SQ- and MQ-LNS reflect impact across the spectrum from efficacy trials to effectiveness studies in a real-world context. These two meta-analyses are also comparable with regard to the age at which supplementation began (generally at 6 mo) and the duration of supplementation (generally 12-18 mo).

It may seem counter-intuitive that a larger quantity of LNS would have less of an impact on linear growth than a smaller quantity. However, a key consideration with regard to efficacy is the actual intake of micronutrients and essential fatty acids from the supplement by the target child. For children in this age range, particularly those < 12 mo of age, the full daily ration of MQ-LNS (220-280 kcal/d) may be too large in relation to energy needs, particularly for 6-11 mo olds who require only ∼200-300 kcal/d from complementary foods. This could lead to incomplete consumption of the supplement and thus a lower than intended intake of micronutrients and fatty acids. There is evidence from several studies that children offered MQ-LNS often consume far less than the daily ration (11–13). As we have demonstrated, if a child consumes only half of a 50g ration of MQ-LNS, the amounts of certain key nutrients (iron, calcium, zinc, vitamin A and thiamin) would fall well short of the recommended intakes. This may not be of concern if the rest of the child’s diet provides those nutrients in adequate amounts, but that is unlikely to be the case for iron, calcium and zinc, which are the most limiting nutrients in complementary food diets (34, 35). As a result, the growth response to MQ-LNS may be less than expected, especially for linear growth.

In conclusion, the current evidence suggests that MQ-LNS offers no added benefits over SQ-LNS for prevention of child undernutrition. One possible explanation is incomplete consumption of the MQ-LNS ration and thus lower than desirable intake of certain key nutrients. Given that MQ-LNS is more expensive per daily ration than SQ-LNS, this has important programmatic implications. Further research is needed, particularly rigorous research on the seasonal use of MQ-LNS, the use of MQ-LNS in highly food insecure settings, and studies directly comparing MQ-LNS vs. SQ-LNS.

## Data Availability

Data described in the manuscript are publicly available via referenced articles and the aggregated data and code are available upon request.

## Acknowledgments

We thank members of the SQ-LNS Task Force for input during the design and implementation of the systematic review and meta-analysis. The authors’ responsibilities were as follows—KGD: drafted the manuscript with input from other coauthors; KRW, CDA, KGD, and CPS: wrote the statistical analysis plan; KRW and CDA: compiled the data; CDA: conducted the data analysis; and all authors: read, contributed to, and approved the final manuscript; KGD is responsible for final content. Supported by Bill & Melinda Gates Foundation grant OPP49817 (to KGD). All authors report no conflicts of interest.

## Abbreviations

GRADE: Grading of Recommendations Assessment, Development and Evaluation

HAZ: height-for-age z-score

IPD: individual participant data

IYCF: infant and young child feeding

LAZ: length-for-age z-score

LNS: lipid-based nutrient supplements

LQ-LNS: large-quantity lipid-based nutrient supplements

MAM: moderate acute malnutrition

MD: mean difference

MQ-LNS: medium-quantity lipid-based nutrient supplements

MUAC: mid-upper arm circumference

MUACZ: mid-upper arm circumference-for-age z-score

PD: prevalence difference

PR: prevalence ratios

RCT: randomized controlled trial

RNI: recommended nutrient intake

RUTF: ready-to-use therapeutic food

SAM: severe acute malnutrition

SBCC: social and behavior change communication

SMC: seasonal malaria chemoprevention

SQ-LNS: small-quantity lipid based nutrient supplements

UCT: unconditional cash transfer

WAZ: weight-for-age z-score

WFP: World Food Programme

WHZ: weight-for-height z-score

WLZ: weight-for-length z-score

WSB+: wheat-soy blend plus Registry and registry number for systematic reviews or meta-analyses: Registered with PROSPERO as CRD42022382448 on December 18, 2022: https://www.crd.york.ac.uk/prospero/display_record.php?ID=CRD42022382448.

---

Supplemental Figure 1: Summary risk of bias as a percentage of all included studies for the effects of MQ-LNS on growth outcomes. MQ-LNS, medium-quantity lipid-based nutrient supplements

Supplemental Figure 2: Forest plot of effect of MQ-LNS vs. control on change in WLZ. MQ-LNS, medium-quantity lipid-based nutrient supplements; WLZ, weight-for-length z-score. Individual study difference-in-difference estimates were extracted from published material and pooled using inverse-variance weighting with random effects. For Adubra (27, 28), values are based on a combined weighted average of the mean change for the SNACK+LNS and SNACK + CASH + LNS groups vs the combined weighted average of the mean change for the SNACK and SNACK + CASH groups. For Soofi (Sindh) (30), values were based on the change in z-scores between 6 and 24 mo (using an approximation of the SD).

Supplemental Figure 3: Forest plot of effect of MQ-LNS vs. control on change in LAZ. MQ-LNS, medium-quantity lipid-based nutrient supplements; LAZ, length-for-age z-score. Individual study difference-in-difference estimates were extracted from published material and pooled using inverse-variance weighting with random effects. For Soofi (Sindh) (30), values were based on the change in z-scores between 6 and 24 mo (using an approximation of the SD).

Supplemental Figure 4: Forest plot of effect of MQ-LNS vs. control on change in WAZ. MQ-LNS, medium-quantity lipid-based nutrient supplements; WAZ, weight-for-age z-score. Individual study difference-in-difference estimates were extracted from published material and pooled using inverse-variance weighting with random effects. For Soofi (Sindh) (30), values were based on the change in z-scores between 6 and 24 mo (using an approximation of the SD).

Supplemental Figure 5: Forest plot of effect of MQ-LNS vs. control on underweight prevalence. MQ-LNS, medium-quantity lipid-based nutrient supplements; PR, prevalence ratio. Individual study prevalence estimates were extracted from published material and prevalence ratios were pooled using inverse-variance weighting with random effects. Pooled estimates were generated using inverse-variance weighting with both fixed and random effects. For Soofi (Sindh) (30), PRs were calculated after recalibrating the endline prevalence differences to take into account the intervention group difference already present at 6 mo, so the effect estimates are related only to the effects of child supplementation.

