## Supplemental Table 2 for "Lipid-based nutrient supplements for prevention of child undernutrition: when less may be more"

**Supplemental Table 2: Risk of bias assessment in each trial**

| Country | Author | Random sequence generation | Allocation concealment | Blinding participants | Blinding outcome assessment^1^ | Incomplete outcome | Selective reporting | Other |
| --- | --- | --- | --- | --- | --- | --- | --- | --- |
| Bangladesh | Christian 2015 | low | low | high | high | low | low | low |
| Malawi | Phuka 2008 | low | low | high | low | low | low | low |
| Malawi | Mangani 2015 | low | low | high | low | low | low | low |
| Malawi | Maleta 2015 | low | low | high | low | low | low | low |
| Mali | Adubra 2019 | low | low | high | high | low | low | low |
| Pakistan - Sindh | Khan 2020 | low | low | high | high | low | low | low |
| Pakistan - Sindh | Soofi 2022 | low | low | high | high | low | low | low |
| Pakistan - Punjab | Soofi 2022 | low | low | high | high | low | low | low |

^1^Due to the nature of the intervention, blinding of participants was not possible. We considered outcome assessment to be at low risk of bias only when it was clearly specified that data collectors who performed the outcome assessments, and the analysts who completed the data analyses, were not aware of group allocation, and it would be unlikely that they could easily become aware of group allocation (i.e., observation of interventions materials in study communities, non-intervention passive control arms, etc.).

| **Christian 2015** |  |  |
| --- | --- | --- |
| **Bias** | **Authors’ judgement** | **Support for judgement** |
| Random sequence generation (selection bias) | Low risk | **Quote:** “A random-number seed was selected by a statistician not involved in the study, using a random number generator, and a random number between 0 and 1 drawn from a uniform distribution was assigned to each sector”  **Comment:** adequately done |
| Allocation concealment (selection bias) | Low risk | **Quote: “**Cluster-randomization of the 596 predefined communities in JiVitA, called ‘sectors’, was done by blocks of 19 (total 32 blocks, last block had 7 sectors). A random-number seed was selected by a statistician not involved in the study, using a random number generator, and a random number between 0 and 1 drawn from a uniform distribution was assigned to each sector. Additionally, a block number was assigned to each sector in groups of 19. For blocks 1–31, the first five sectors by sort order were assigned to treatment group 1, the next five to treatment group 2, and so on. For block 32, the two larger controls were assigned two sectors and the intervention groups 1 sector each.  **Comment**: central randomization of a cluster-randomized trial |
| Blinding of participants and personnel (performance bias) | High risk | **Quote:** “Our trial was unblinded”  **Comment:** not done |
| Blinding of outcome assessment (detection bias) | High risk | **Quote:** “Our trial was unblinded”; “Anthropometry was done at this time using standard methods…Repeat infant anthropometry assessments were conducted during home visits every 3 months after the baseline at 9, 12, 15 and 18 months of age.”  **Comment**: not done; cluster-randomized trial |
| Incomplete outcome data (attrition bias) | Low risk | **Attrition:** control group = 1312/1591; Plumpy’Doz group = 1395/1599; rice lentil group = 785/901; chickpea group = 786/920; WSB++ group = 789/928  **Comments**: reasons given for loss to follow-up; missing outcome data balanced in numbers across intervention groups |
| Selective reporting (reporting bias) | Low risk | **Comment:** trial registered as NCT01562379 at ClinicalTrials.gov, outcomes described in the methodology section reported in the results section |
| Other bias | Low risk | **Comment:** no other potential sources of bias reported |

| **Phuka 2008** |  |  |
| --- | --- | --- |
| **Bias** | **Authors’ judgement** | **Support for judgement** |
| Random sequence generation (selection bias) | Low risk | **Quote:** “The randomization list and envelopes were made by individuals not involved in trial implementation”  **Comment:** adequately done |
| Allocation concealment (selection bias) | Low risk | **Quote:** “For group allocation, guardians chose 1 envelope from a set of identical appearing opaque envelopes, each containing a piece of paper indicating an identification number and randomly assigned allocation to 1 of the 3 interventions”.  **Comment**: adequately done |
| Blinding of participants and personnel (performance bias) | High risk | **Comment**: participant blinding not possible due to the nature of the intervention (LNS, FBF, Control) |
| Blinding of outcome assessment (detection bias) | Low risk | **Quote:** “the people measuring the outcomes were masked to group allocation”  **Comment:** adequately done |
| Incomplete outcome data (attrition bias) | Low risk | **Attrition**: LP (FBF) = 57/61; FS50 (LNS) = 54/61; FS25 (LNS) = 57/60  **Comment:** reasons given for loss to follow-up; missing outcome data balanced in numbers across intervention groups |
| Selective reporting (reporting bias) | Low risk | **Comment:** The trial was registered at ClinicalTrials.gov (NCT00131209); outcomes described in the methods section reported in the results section |
| Other bias | Low risk | **Comment:** no other potential sources of bias reported |

| **Mangani 2015** |  |  |
| --- | --- | --- |
| **Bias** | **Authors’ judgement** | **Support for judgement** |
| Random sequence generation (selection bias) | Low risk | **Quote:** “Blocked randomization, with each block containing 16 allocations evenly distributed for the four groups, was used to assign participants to intervention groups…The randomization list and envelopes were made by an individual not involved in trial implementation.”  **Comment**: adequately done |
| Allocation concealment (selection bias) | Low risk | **Quote**: “A set of identical-appearing opaque envelopes from one randomization block was shuffled and a guardian was requested to choose one envelope. The envelope contained an identification number and the allocation to one of the four interventions.”  **Comment:** adequately done |
| Blinding of participants and personnel (performance bias) | High risk | **Comment:** participant blinding not possible due to the nature of the intervention (LNS, CSB, Control) |
| Blinding of outcome assessment (detection bias) | Low risk | **Quote:** “The internal validity of the trial was high because of … blinding of the outcome assessors”  **Comment**: adequately done |
| Incomplete outcome data (attrition bias) | Low risk | **Attrition:** Control group = 185/209; Milk-LNS group = 191/212; Soy-LNS group = 188/210; CSB = 183/209  **Comment:** Minimal attrition and reasons given for loss to follow-up; missing outcome data balanced in numbers across intervention groups |
| Selective reporting (reporting bias) | Low risk | **Comment**: The trial was registered at clinicaltrials.gov (NCT00524446); outcomes described in the methods section reported in the results section |
| Other bias | Low risk | **Comment:** no other potential sources of bias reported |

| **Maleta 2015** |  |  |
| --- | --- | --- |
| **Bias** | **Authors’ judgement** | **Support for judgement** |
| Random sequence generation (selection bias) | Low risk | **Quote:** “We used block randomization and a set of opaque envelopes to assign participants to the intervention groups. The randomization list and envelopes were prepared by a study statistician not involved in trial implementation”  **Comment:** adequately done |
| Allocation concealment (selection bias) | Low risk | **Quote:** “We used block randomization and a set of opaque envelopes to assign participants to the intervention groups.”  **Comment:** adequately done |
| Blinding of participants and personnel (performance bias) | High risk | **Comment**: participant blinding not possible due to the nature of the intervention (LNS, Control) |
| Blinding of outcome assessment (detection bias) | Low risk | **Quote:** “…and the code was not disclosed to the researchers or to those assessing the outcomes until all data had been entered and verified in a database.” “For the LNS group, we used single-masked procedures (i.e., fieldworkers who delivered the supplements knew which children were receiving LNSs, but those who performed the anthropometric measurements or assessed other outcomes were not aware of group allocation).”  **Comment:** adequately done |
| Incomplete outcome data (attrition bias) | Low risk | **Attrition:** 40g/day milk-free LNS group =239/324; 40g/day milk LNS group = 242/322; 20g/day milk-free LNS group = 247/323; 20g/day milk LNS group = 236/322;10g/day milk LNS group = 221/321; control group = 242/320  **Comment**: similar levels of attrition across groups, reasons for dropout provided |
| Selective reporting (reporting bias) | Low risk | **Comment**: The trial was registered at clinicaltrials.gov (NCT00945698); SAP and trial protocol available online; outcomes described in the methods section reported in the results section |
| Other bias | Low risk | **Comment:** no other potential sources of bias reported |

| **Adubra 2019** |  |  |
| --- | --- | --- |
| **Bias** | **Authors’ judgement** | **Support for judgement** |
| Random sequence generation (selection bias) | Low risk | **Quote**: “to minimize disparities between CHCs, they were matched to make blocks of 4 in each district…during a public event, the representatives of the CHCs in each block of 4 were asked to draw blindly and successively a ball from a bag containing 4 different colored balls. Each color was then assigned a treatment using a table of random numbers with Emergency Nutrition Assessment software”  **Comment:** adequately done |
| Allocation concealment (selection bias) | Low risk | **Quote**: “to minimize disparities between CHCs, they were matched to make blocks of 4 in each district…during a public event, the representatives of the CHCs in each block of 4 were asked to draw blindly and successively a ball from a bag containing 4 different colored balls. Each color was then assigned a treatment using a table of random numbers with Emergency Nutrition Assessment software”  **Comment:** central randomization of a cluster-randomized controlled trial |
| Blinding of participants and personnel (performance bias) | High risk | **Quote**: “program participants and individuals delivering the interventions were not masked to cluster assignment””  **Comment**: blinding of participants who received no intervention was not possible |
| Blinding of outcome assessment (detection bias) | High risk | **Quote:** “enumerators were not told the allocation of the participants they were interviewing; however, at endline, the questionnaire included questions about whether mothers and their children received cash and/or LNS. Enumerators performing anthropometric measurements were masked. [Authors] were not masked when doing the analysis”  **Comment**: analysts not masked |
| Incomplete outcome data (attrition bias) | Low risk | **Quote**: “Two repeated cross-sectional surveys were carried out on independent representative samples…before the start of the cash and LNS components…and 3 years later”.  **Comment**: two repeated cross-sectional surveys, not applicable |
| Selective reporting (reporting bias) | Low risk | **Comment**: trial registered as ISRCTN08435964 ([www.isrctn.com](http://www.isrctn.com)); outcomes described in the methods section reported in the results section |
| Other bias | Low risk | **Comment**: no other potential sources of bias reported |

| **Khan 2020 (Sindh, Pakistan Cohort 2)** |  |  |
| --- | --- | --- |
| **Bias** | **Authors’ judgement** | **Support for judgement** |
| Random sequence generation (selection bias) | Low risk | **Quote:** “Of the 29 UCs [union councils], where the stunting prevention programme was implemented, 12 UCs were randomly allocated to intervention and control groups with a computer-generated randomization sequence that was generated by an independent expert at the Data Management Unit in Agha Khan University”  **Comment:** adequately done |
| Allocation concealment (selection bias) | Low risk | **Quote:** “Of the 29 UCs [union councils], where the stunting prevention programme was implemented, 12 UCs were randomly allocated to intervention and control groups with a computer-generated randomization sequence that was generated by an independent expert at the Data Management Unit in Agha Khan University”  **Comment:** central randomization of a cluster-randomized controlled trial |
| Blinding of participants and personnel (performance bias) | High risk | **Comment**: participant blinding not possible due to the nature of the intervention (LNS, Control) |
| Blinding of outcome assessment (detection bias) | High risk | **Quote:** “Data on compliance was collected using parental recall and observation of used and unused sachets at household level. Anthropometric data was collected on a quarterly basis”.  **Comment:** outcome assessment done by an independent team (different from those distributing supplements); however, outcome assessors also assessed compliance with the intervention |
| Incomplete outcome data (attrition bias) | Low risk | **Attrition:** “A total of 428 (94.9%) children in the control group and 402 (95.9%) children in the intervention group completed all their follow-up visits by 24 months of age or had follow-up assessments for at least six months”  **Comment:** reasons for loss-to-follow-up mentioned; missing outcome data balanced in numbers across intervention groups |
| Selective reporting (reporting bias) | Low risk | **Comment**: Trial registered at ClinicalTrials.gov (NCT02422953); outcomes described in the methodology section reported in the results section |
| Other bias | Low risk | **Comment**: no other potential sources of bias reported |

| **Soofi 2022 (Sindh, Pakistan Cohort 1)** |  |  |
| --- | --- | --- |
| **Bias** | **Authors’ judgement** | **Support for judgement** |
| Random sequence generation (selection bias) | Low risk | **Quote:** “Out of 29 stunting prevention program UCs, 12 UCs were randomly selected and equally assigned to the intervention or control groups through a computer-generated randomization sequence by the data management unit at the Aga Khan University”  **Comment:** adequately done |
| Allocation concealment (selection bias) | Low risk | **Quote:** “Out of 29 stunting prevention program UCs, 12 UCs were randomly selected and equally assigned to the intervention or control groups through a computer-generated randomization sequence by the data management unit at the Aga Khan University”  **Comment:** central randomization of a cluster-randomized controlled trial |
| Blinding of participants and personnel (performance bias) | High risk | **Comment**: participant blinding not possible due to the nature of the intervention (LNS, Control) |
| Blinding of outcome assessment (detection bias) | High risk | **Quote:** “The intervention package included a distinctly visible component; therefore, they study participants and data collection team were not masked in the intervention assignment. However, data collection teams were different in both groups, and data analysts remained blinded to study arms until the final analysis was completed”.  **Comment:** data collection teams not blinded |
| Incomplete outcome data (attrition bias) | Low risk | **Attrition:** Among the children available at 6 months: Control = 653/705; LNS = 699/753  **Comment:** reasons for loss-to-follow-up mentioned; missing outcome data balanced in numbers across intervention groups |
| Selective reporting (reporting bias) | Low risk | **Comment**: Trial registered at ClinicalTrials.gov (NCT02422953); outcomes described in the methodology section reported in the results section |
| Other bias | Low risk | **Comment**: no other potential sources of bias reported |

| **Soofi 2022 (Punjab)** |  |  |
| --- | --- | --- |
| **Bias** | **Authors’ judgement** | **Support for judgement** |
| Random sequence generation (selection bias) | Low risk | **Quote:** “A 2-stage stratified random sampling strategy was used to minimize the risk of contamination among study arms…a total of 200 clusters were randomly selected and assigned into 1 of 4 study arms…Randomization was conducted by an independent statistician who was not involved in the study”  **Comment:** adequately done |
| Allocation concealment (selection bias) | Low risk | **Quote:** “A 2-stage stratified random sampling strategy was used to minimize the risk of contamination among study arms…a total of 200 clusters were randomly selected and assigned into 1 of 4 study arms…Randomization was conducted by an independent statistician who was not involved in the study”  **Comment**: central randomization of a cluster-randomized trial |
| Blinding of participants and personnel (performance bias) | High risk | **Quote:** “The blinding of study participants and study arms was not possible from data collection teams and investigators as they were responsible for supervising the provision of LNS-MQ and SBCC sessions”  **Comment:** blinding of participants and personnel was not possible due to the nature of the intervention |
| Blinding of outcome assessment (detection bias) | High risk | **Quote**: “The blinding of study participants and study arms was not possible from data collection teams and investigators as they were responsible for supervising the provision of LNS-MQ and SBCC sessions. However, data analysts remained blinded to study participants and study arms until the final data analysis was completed”  **Comment:** data collection teams not blinded |
| Incomplete outcome data (attrition bias) | Low risk | **Attrition**: UCT = 410/434; UCT + SBCC = 425/433; UCT + LNS = 421/430; UCT + LNS+SBCC = 419/432  **Comment:** reasons for loss-to-follow-up mentioned; missing outcome data balanced in numbers across intervention groups |
| Selective reporting (reporting bias) | Low risk | **Comment: T**rial registered as  NCT03299218 at ClincialTrials.gov.; outcomes described in the methods section reported in the results section |
| Other bias | Low risk | **Comment:** no other potential sources of bias reported |
