## Supplemental Figure 1 for "Lipid-based nutrient supplements for prevention of child undernutrition: when less may be more"

### Slide 1
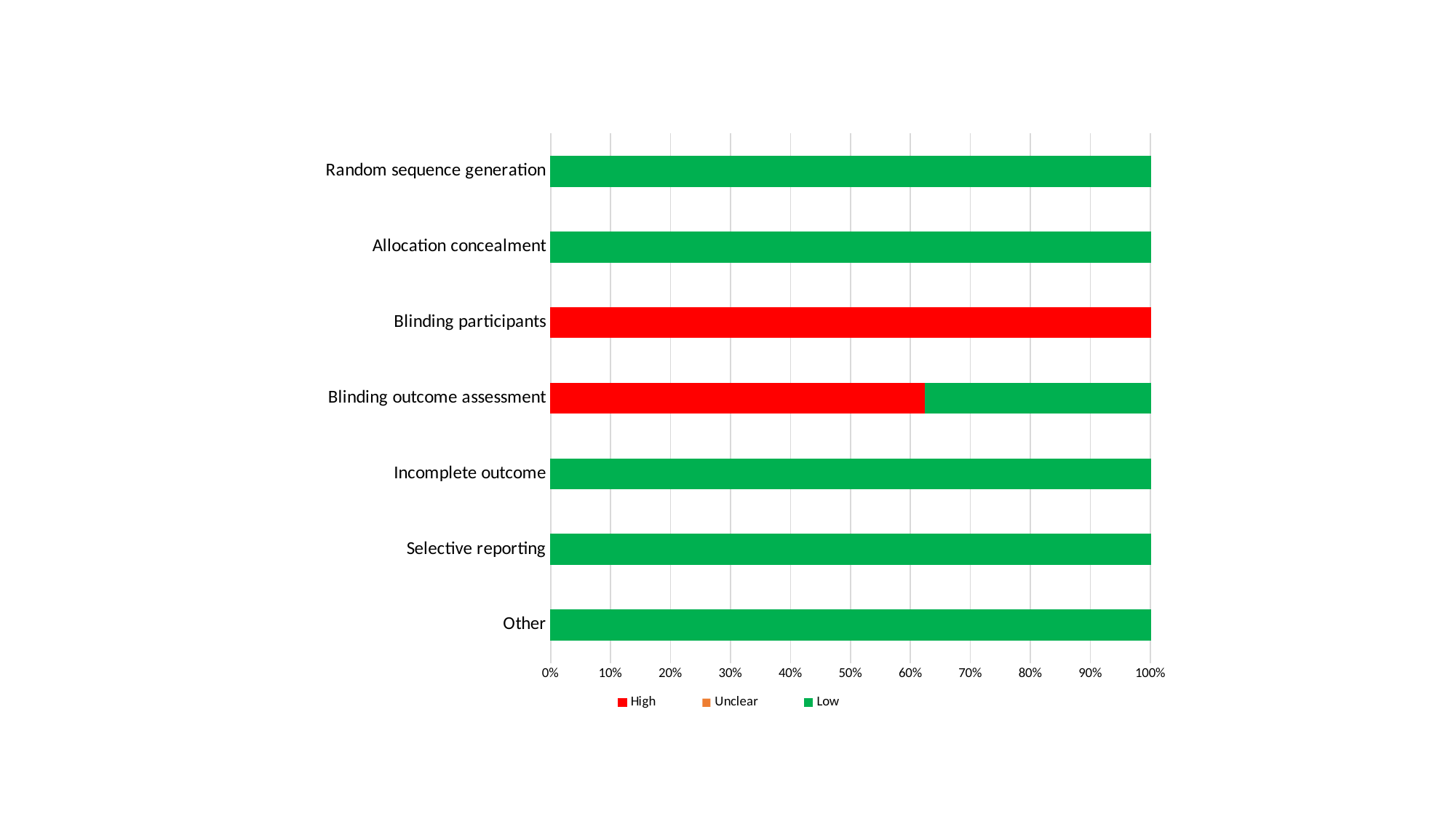

#### Chart
| Category | High | Unclear | Low |
|---|---|---|---|
| Random sequence generation | 0.0 | 0.0 | 1.0 |
| Allocation concealment | 0.0 | 0.0 | 1.0 |
| Blinding participants | 1.0 | 0.0 | 0.0 |
| Blinding outcome assessment | 0.625 | 0.0 | 0.375 |
| Incomplete outcome | 0.0 | 0.0 | 1.0 |
| Selective reporting | 0.0 | 0.0 | 1.0 |
| Other | 0.0 | 0.0 | 1.0 |
