## Supplementary figures and images for "Lipid-based nutrient supplements for prevention of child undernutrition: when less may be more"

### Supplemental Figure 2

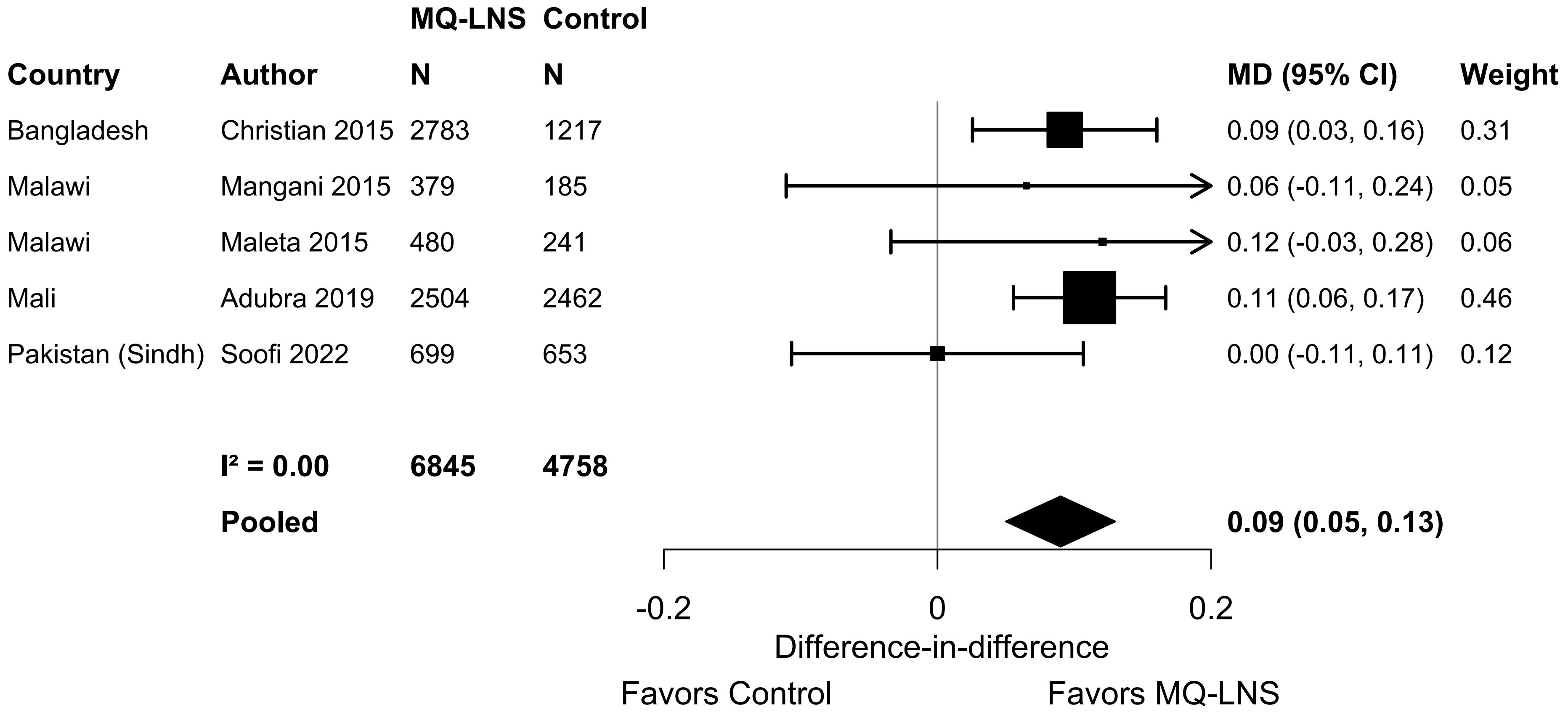

### Supplemental Figure 3

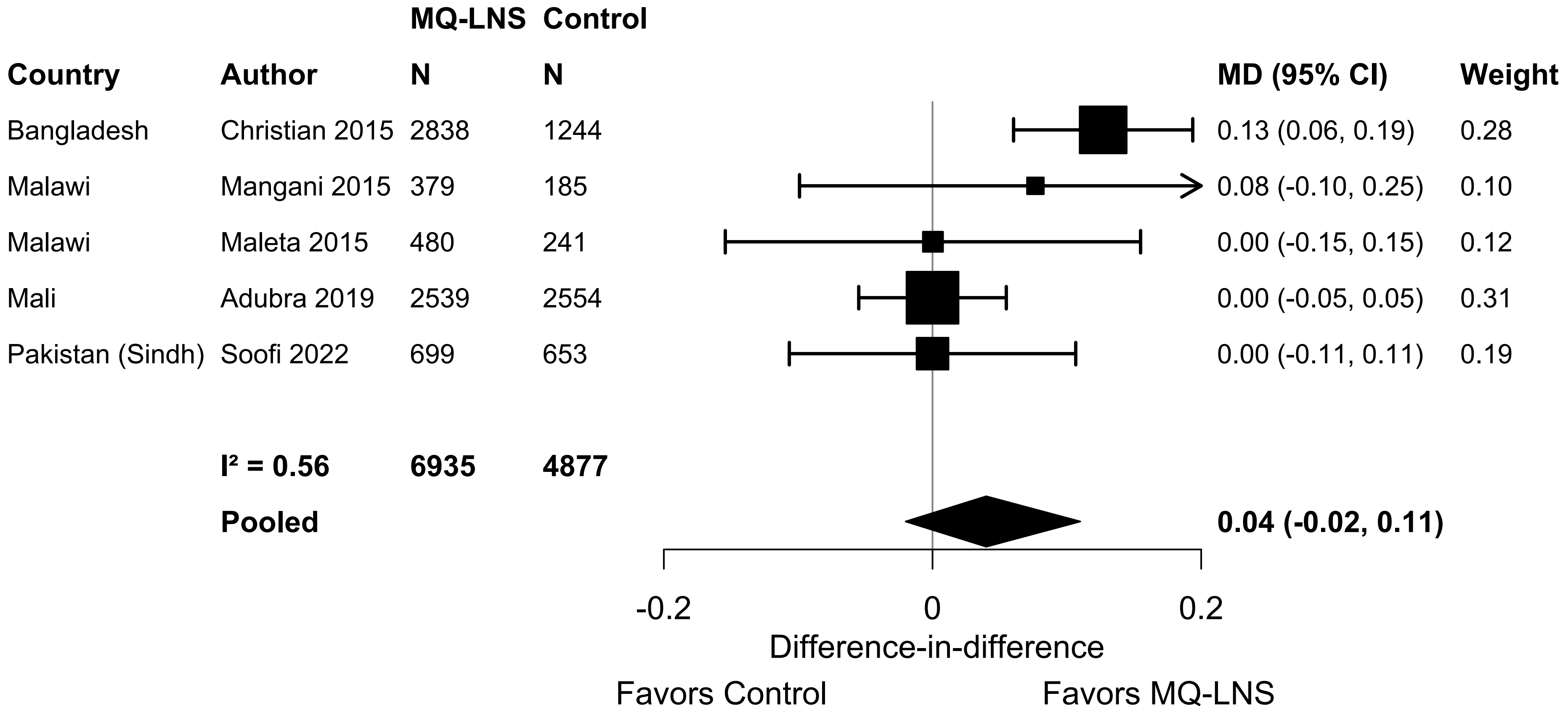

### Supplemental Figure 4

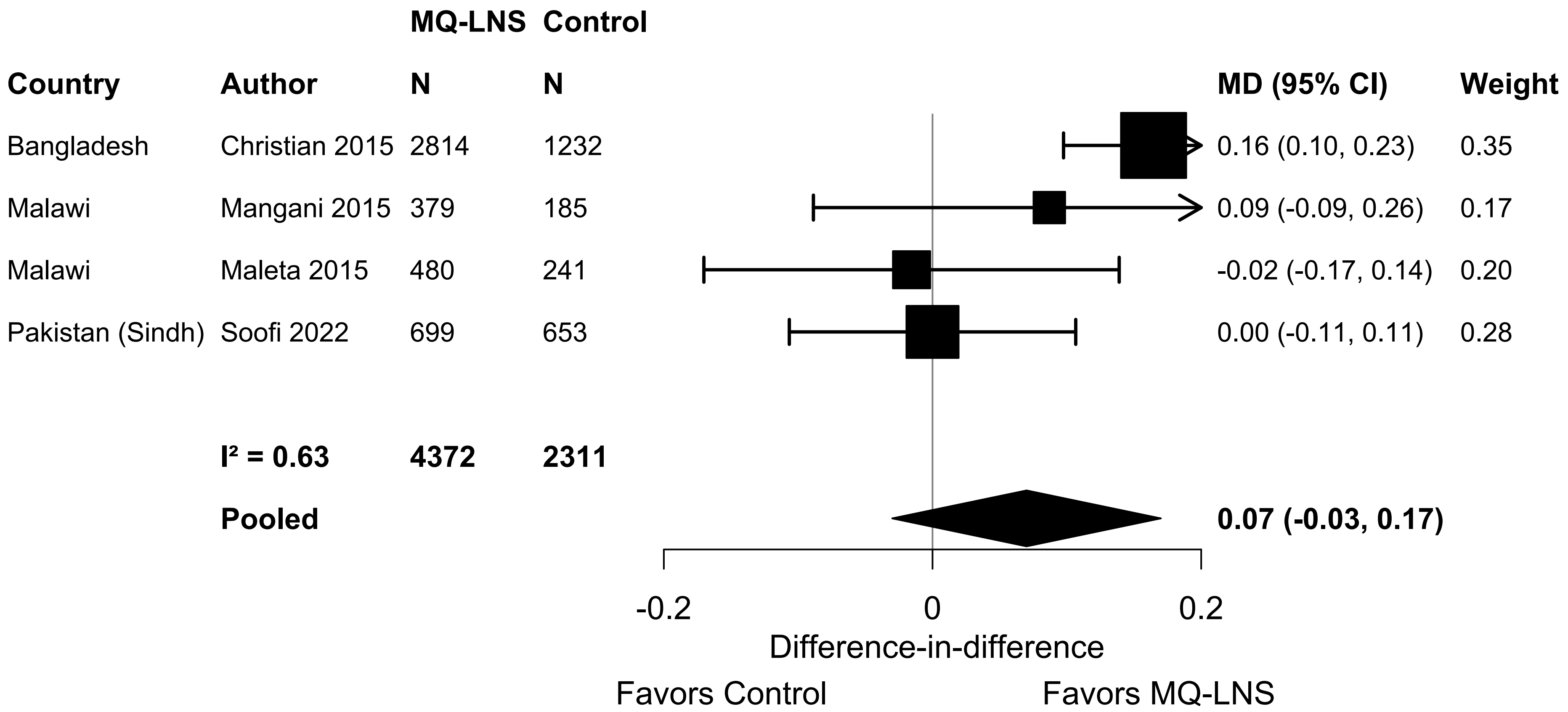
