## Supplemental Figure 5 for "Lipid-based nutrient supplements for prevention of child undernutrition: when less may be more"

| Country | Author | MQ-LNS N | Control N | PR (95% CI) | Weight |
|---|---|---|---|---|---|
| Bangladesh | Christian 2015 | 2839 | 1251 | 0.89 (0.81, 0.96) | 0.40 |
| Malawi | Maleta 2015 | 474 | 251 | 2.12 (1.20, 3.73) | 0.01 |
| Pakistan (Sindh) | Khan 2020 | 351 | 357 | 0.94 (0.81, 1.10) | 0.14 |
| Pakistan (Sindh) | Soofi 2022 | 699 | 653 | 0.96 (0.87, 1.05) | 0.35 |
| Pakistan (Punjab) | Soofi 2022 (SBCC) | 419 | 425 | 0.98 (0.76, 1.27) | 0.05 |
| Pakistan (Punjab) | Soofi 2022 (no SBCC) | 421 | 410 | 0.95 (0.73, 1.24) | 0.05 |
| | **I² = 0.06** | **5203** | **3347** | | |
| | **Pooled** | | | **0.94 (0.88, 0.99)** | |

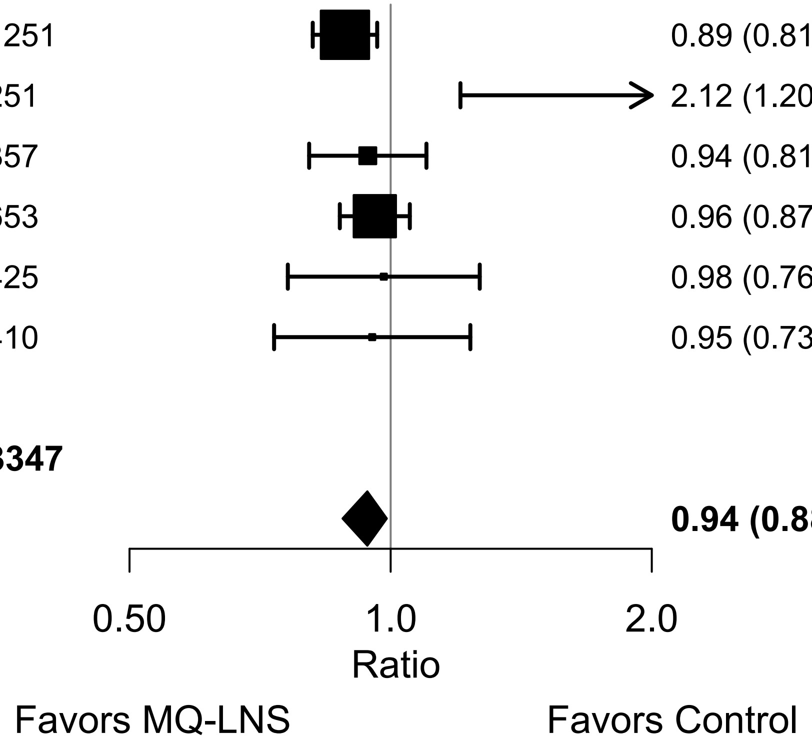
